# Cortical Lesions Form Predominantly in Early Multiple Sclerosis

**DOI:** 10.64898/2026.04.30.26352141

**Authors:** Batuhan Ayci, Emma Dereskewicz, Jonadab Dos Santos Silva, Julia Galasso, Phoebe Rust, Francesco La Rosa, Jiaen Liu, Daniel S Reich, James F Sumowski, Erin S Beck

## Abstract

**Background and Objectives:** Cortical lesions are common in multiple sclerosis (MS) and associated with disability, but their characterization in early MS has been limited. Here, we aimed to characterize cortical lesions in newly diagnosed MS with 7 tesla (T) brain MRI.

**Methods:** Adults within 14 months of relapsing-remitting MS diagnosis underwent 7T brain MRI and clinical evaluation at Mount Sinai. Cortical lesions were identified using T1-weighted (w) (median of three acquisitions) and T2*w images (both at 0.5mm^3^). Non-cortical brain lesions were segmented on 0.7mm^3^ T1w images. Lesion burden in newly diagnosed MS was compared with a previously analyzed NIH cohort with longer time since diagnosis, imaged using a similar protocol.

**Results:** 61 individuals were included in the newly diagnosed MS cohort (mean age 34 ± 4 years; 72% female; median time since diagnosis 5 months, interquartile range [IQR] 6). Cortical lesions were identified in 50/61 (81%) individuals, and subpial cortical lesions were identified in 46 (75%). Median cortical lesion number was 5 (IQR 11), median volume 319 μl (IQR 1049). Cortical lesions constituted a median of 14% of total brain lesion volume (IQR 43%), and in 21% of individuals, cortical lesions constituted >50% of total brain lesion volume. Cortical lesion number was associated with worse 9-hole peg test (ρ=0.33, p=0.008) and Symbol Digit Modalities Test performance (ρ=-0.29, p=0.02). When pooled with the NIH cohort (n=60, median time since diagnosis 12 years, IQR 17), non-cortical lesion volume was ∼3.5 times higher in people with time since diagnosis >36 months (median 4.7 ml, IQR 8.7) vs ≤36 months (median 1.2 ml, IQR 2.4, p<0.001). In contrast, cortical lesion volume was only ∼1.3 times higher in people with time since diagnosis >36 months (median 416 μl, IQR 1013) vs ≤36 months (median 318 μl, IQR 925, p=0.04). Non-cortical lesion volume was moderately associated with time since diagnosis (ρ=0.54, p<0.001) vs ρ=0.27 (p<0.001) for cortical lesions.

**Discussion:** Cortical lesions are prevalent in newly diagnosed MS and constitute a substantial portion of total lesion burden. Cortical lesion volume is similar in early vs established MS, suggesting most cortical lesions form early in disease.

## Introduction

Multiple sclerosis (MS) is a chronic immune-mediated demyelinating disease of the central nervous system, affecting both white and gray matter. While focal white matter demyelination is the radiological hallmark of the disease, histopathological studies have long demonstrated that cortical demyelination is common and can be extensive^1–4^. However, cortical lesions remain largely undetected on conventional MRI due to their small size and low contrast vs surrounding gray matter. Advances in MRI have improved the detection of cortical pathology, particularly at ultra-high field strength^5–7^. Combined use of high-resolution, optimized T1-weighted (T1w) and T2*-weighted (T2*w) MRI sequences at 7 tesla (T) greatly improves visualization of cortical lesions, particularly subpial lesions that are nearly invisible at lower field strengths^8–11^.

Cortical lesions have been associated with disability accumulation and progressive disease, with evidence suggesting a greater clinical relevance than white matter lesions^12,13^. However, most existing data on cortical lesions derive mainly from people with longstanding disease, limiting insight into when cortical lesions first form and how early they influence neurological function. Here, we characterized cortical lesions and examined their associations with disability in a newly diagnosed MS cohort.

## Methods

### Clinical cohorts

Participants were enrolled in an Institutional Review Board-approved, prospective, observational study at Icahn School of Medicine at Mount Sinai between 2022 and 2025. All participants provided written informed consent. Inclusion criteria were (1) age 18 years or older, (2) diagnosis of relapsing-remitting MS according to the 2017 McDonald criteria^14^ within the preceding 14 months, and (3) no contraindications to 7 T MRI. Participants underwent clinical evaluation, including Expanded Disability Status Scale (EDSS, n=58), Symbol Digit Modalities Test (SDMT, n=56), timed 25-foot walk (T25FW, n=59), and 9-hole peg test (9HPT, n=61). The SDMT was administered in oral form.

To compare cortical lesion burden between early and established disease, we analyzed data from a second cohort with available 7 T MRI data, acquired at the NIH^15^. In the NIH cohort, the SDMT was administered in written form.

### MRI acquisition

Participants at Mount Sinai and the NIH underwent 7 T brain MRI scans on 7 T whole-body research MRI systems (Siemens, Erlangen, Germany), equipped with a single-channel transmit and 32-channel receive phased array head coil (Nova Medical, Wilmington, MA, USA). Scans at both sites included 3D magnetization prepared 2 rapid gradient echo (MP2RAGE) acquired at 0.5 mm isotropic (acquired three times per session at Mount Sinai and four times per session at the NIH) and 0.7 mm isotropic (acquired once per session) resolution. In addition, at Mount Sinai, motion- and B_0_-corrected 3D T2*w gradient recalled echo (GRE, 0.5 mm isotropic) was acquired in three partially overlapping volumes. Correction for rigid-body head motion and spatially linear B_0_ changes was performed offline^10,16^. At the NIH, 3D echo-planar imaging (EPI; 0.5 mm isotropic, acquired in two partially overlapping volumes), multi-echo T2*w GRE (0.5 mm isotropic, acquired in three partially overlapping volumes) were acquired. Detailed sequence parameters are included in Supplementary Table 1.

At both sites, uniform-denoised MP2RAGE images (hereafter T1w MP2RAGE) were generated using the vendor-provided reconstruction.

### Image processing

The denoised 7 T 0.5 mm T1w MP2RAGE images from each acquisition were co-registered, and a median image was generated, as described previously^15^. The 7 T T2*w GRE magnitude images were calculated as the root mean square of the magnitude across echoes. The averaged T2*w images were rigidly registered to the 0.5 mm T1w MP2RAGE median images using the Advanced Normalization Tools (ANTs) registration framework^17^.

### Lesion segmentation

For the Mount Sinai newly diagnosed MS cohort, cortical lesions were manually segmented using median 0.5 mm T1w MP2RAGE and T2*w GRE using ITK-Snap^18^. Cortical lesions were segmented independently by two raters, followed by consensus review by a third rater, as previously described^15^. Lesions were categorized as leukocortical (involving cortex and white matter and not touching the pial surface of the brain), intracortical (confined to the cortex and not touching the pial surface of the brain), or subpial (involving the cortex and touching the pial surface, with or without white matter involvement)^19^.

For the NIH cohort, cortical lesions were previously segmented using 7 T T1w and T2*w images^15^. To determine rater-related differences in cortical lesion segmentation, a subset of 10 NIH cases was re-segmented by the Mount Sinai raters on 7 T T1w and T2*w images.

Non-cortical lesions, which included lesions in the cerebral white matter, deep gray matter, and infratentorium, were segmented on 0.7 mm T1w MP2RAGE images from both sites using the automated method Pseudo-Label Assisted nnU-Net (PLaN),^20^ followed by manual review and correction. In four cases in the Mount Sinai newly diagnosed MS cohort and three cases in the NIH cohort, for which 0.7 mm MP2RAGE images were unavailable, 0.5 mm MP2RAGE images alone were used.

### Brain volume measurement

Brain volumetrics and cortical thickness in the Mount Sinai cohort were derived from the 0.7 mm T1w MP2RAGE images using Freesurfer’s *recon-all* pipeline (version 7.4.1; http://surfer.nmr.mgh.harvard.edu) for all individuals in which 0.7 mm MP2RAGE was acquired (57/61). First, a lesion-filling preprocessing step was performed using the FMRIB Software Library tool *lesion_filling*^21^. Whole brain, cortex, white matter, and deep grey matter volumes were extracted using SynthSeg^20^. Brain volumes were normalized for head size by dividing each FreeSurfer-derived volume^21^ by the subject’s estimated total intracranial volume (eTIV) and multiplying by the MNI152 reference eTIV^23^.

### Statistical analyses

Inter-rater reliability for cortical lesion segmentation was assessed using the intraclass correlation coefficient, using both subjects and raters as random effects. Associations between MRI and clinical variables were assessed using Spearman’s rank coefficients. Pairwise Wilcoxon signed-rank tests were used to compare groups. Effect sizes were estimated using the Hodges-Lehmann estimator. Sex differences were assessed using Fisher’s exact test. Benjamini-Hochberg false discovery rate was used to correct for multiple comparisons. Statistical significance was set at p<0.05. All analyses were performed in R (version 4.5.1; R Foundation for Statistical Computing, Vienna, Austria) using RStudio (version 2025.05.1+513).

## Results

### Cortical lesions are prevalent in newly diagnosed MS and constitute a substantial portion of total brain lesion volume

61 people (mean age 34 ± 7 years; 72% female) with relapsing-remitting MS within 14 months of diagnosis underwent clinical evaluation and 7 T brain MRI at Mount Sinai (Table 1). The median time since symptom onset was 12 months (interquartile range [IQR] 16, range 0-203), the median time since diagnosis was 5 months (IQR 6, range 0-14), and the median EDSS was 2 (IQR 2, range 0-3).

**Table 1.**
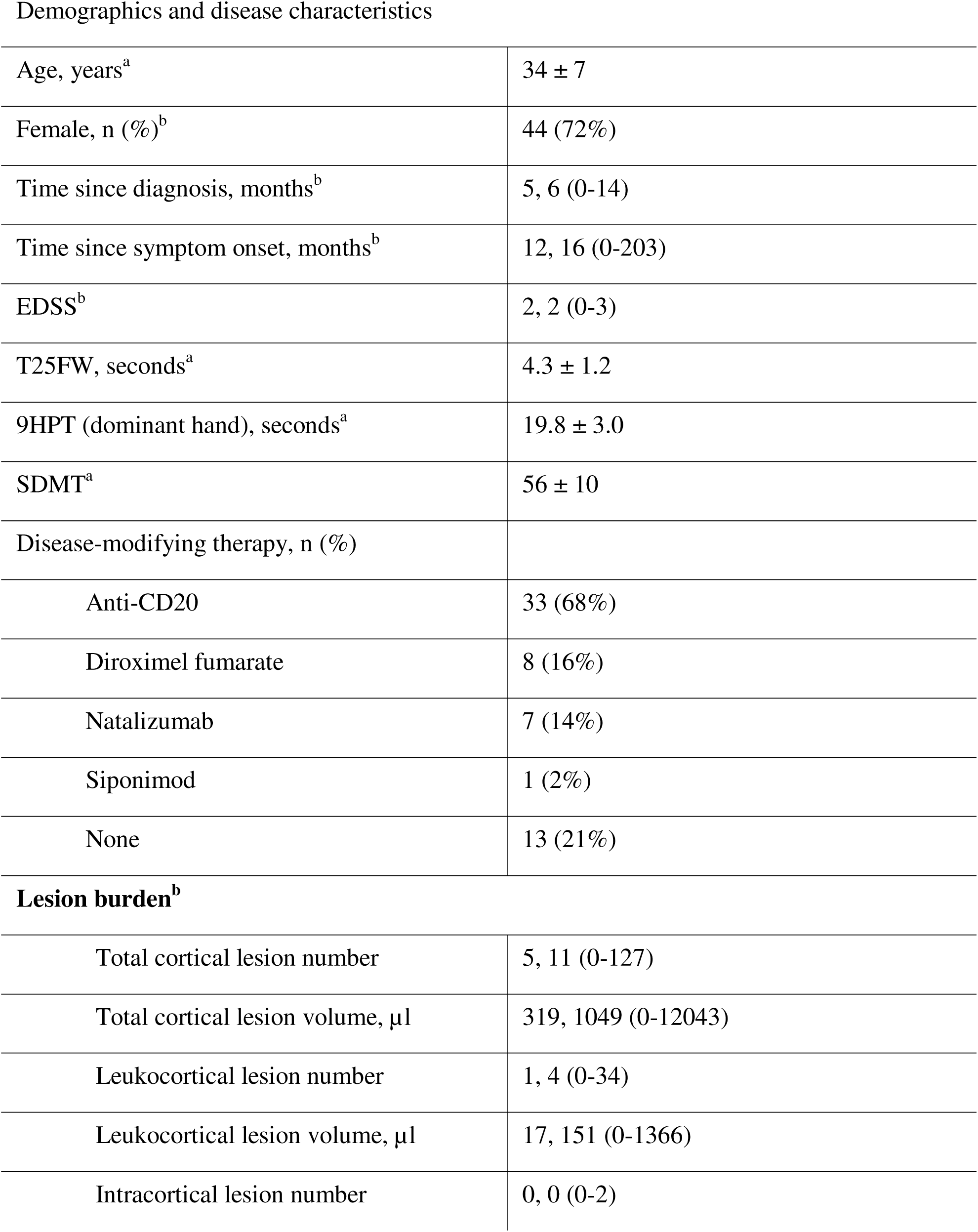
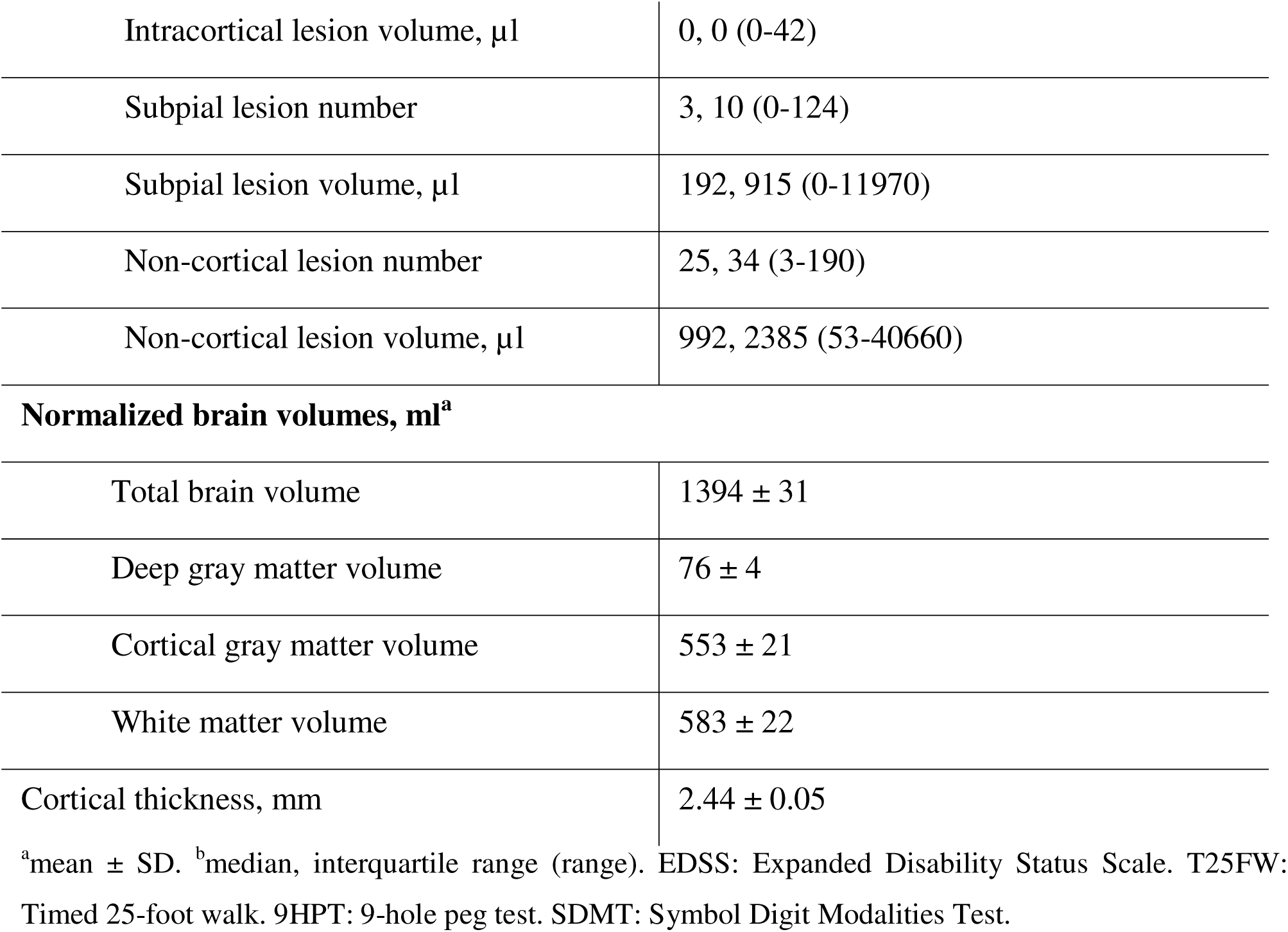
Newly diagnosed relapsing-remitting multiple sclerosis cohort (n=61) demographics, disease characteristics, lesion burden, and brain volumetrics.

|  |  |
| --- | --- |
| Age, years<sup>a</sup> | 34 ± 7 |
| Female, n (%)<sup>b</sup> | 44 (72%) |
| Time since diagnosis, months<sup>b</sup> | 5, 6 (0-14) |
| Time since symptom onset, months<sup>b</sup> | 12, 16 (0-203) |
| EDSS<sup>b</sup> | 2, 2 (0-3) |
| T25FW, seconds<sup>a</sup> | 4.3 ± 1.2 |
| 9HPT (dominant hand), seconds<sup>a</sup> | 19.8 ± 3.0 |
| SDMT<sup>a</sup> | 56 ± 10 |
| Disease-modifying therapy, n (%) |  |
| Anti-CD20 | 33 (68%) |
| Diroximel fumarate | 8 (16%) |
| Natalizumab | 7 (14%) |
| Siponimod | 1 (2%) |
| None | 13 (21%) |
| **Lesion burden<sup>b</sup>** |  |
| Total cortical lesion number | 5, 11 (0-127) |
| Total cortical lesion volume, µl | 319, 1049 (0-12043) |
| Leukocortical lesion number | 1, 4 (0-34) |
| Leukocortical lesion volume, µl | 17, 151 (0-1366) |
| Intracortical lesion number | 0, 0 (0-2) |
| Intracortical lesion volume, $\mu\text{l}$ | 0, 0 (0-42) |
| Subpial lesion number | 3, 10 (0-124) |
| Subpial lesion volume, $\mu\text{l}$ | 192, 915 (0-11970) |
| Non-cortical lesion number | 25, 34 (3-190) |
| Non-cortical lesion volume, $\mu\text{l}$ | 992, 2385 (53-40660) |
| **Normalized brain volumes, $\text{ml}^{\text{a}}$** |  |
| Total brain volume | $1394 \pm 31$ |
| Deep gray matter volume | $76 \pm 4$ |
| Cortical gray matter volume | $553 \pm 21$ |
| White matter volume | $583 \pm 22$ |
| Cortical thickness, mm | $2.44 \pm 0.05$ |

<sup>a</sup>mean ± SD. <sup>b</sup>median, interquartile range (range). EDSS: Expanded Disability Status Scale. T25FW: Timed 25-foot walk. 9HPT: 9-hole peg test. SDMT: Symbol Digit Modalities Test.

A total of 704 cortical lesions were identified (median per participant 5, IQR 15, range 0-127), including 169 leukocortical lesions (median 1, IQR 4, range 0-34), 13 intracortical lesions (median 0, IQR 0, range 0-2), and 522 subpial lesions (median 3, IQR 10, range 0-124) (Table 1, Figure 1). 50/61 individuals (82%) had at least one cortical lesion, 33 (54%) had at least one leukocortical lesion, 12 (20%) had at least one intracortical lesion, and 46 (75%) had at least one subpial lesion (Figure 2A). Intraclass correlation coefficient for cortical lesion number between the two raters was 0.89 (95% Confidence Interval [CI]: 0.80-0.94, p<0.001).

**Figure 1.**
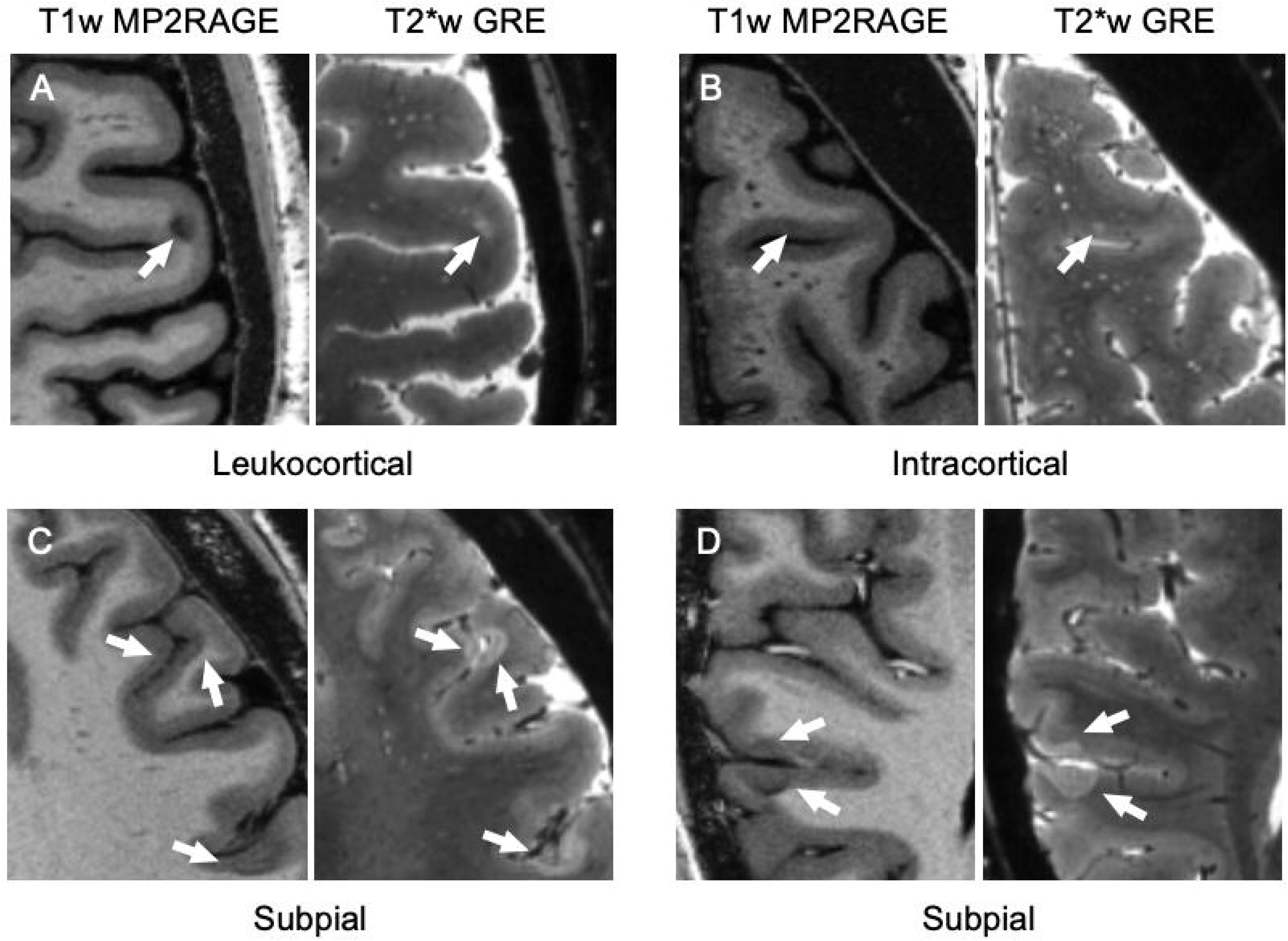
Representative images of cortical lesions. Cortical lesions are indicated by arrows on T1 and T2*-weighted (w) images. Lesions are hypointense on T1w (right) and hyperintense on T2*w (left) images. **(A)** A leukocortical lesion involving both the cortex and the underlying white matter but not extending to the pial surface. **(B)** An intracortical lesion confined entirely to the gray matter, sparing the pial surface and not involving the adjacent white matter. **(C)** A region exhibiting three subpial lesions. **(D)** An example of two subpial lesions on opposite sides of a sulcus. The subpial lesion toward the bottom of the image extends into the white matter. MP2RAGE: magnetization prepared 2 rapid gradient echoes. GRE: gradient-recalled echo.

**Figure 2.**
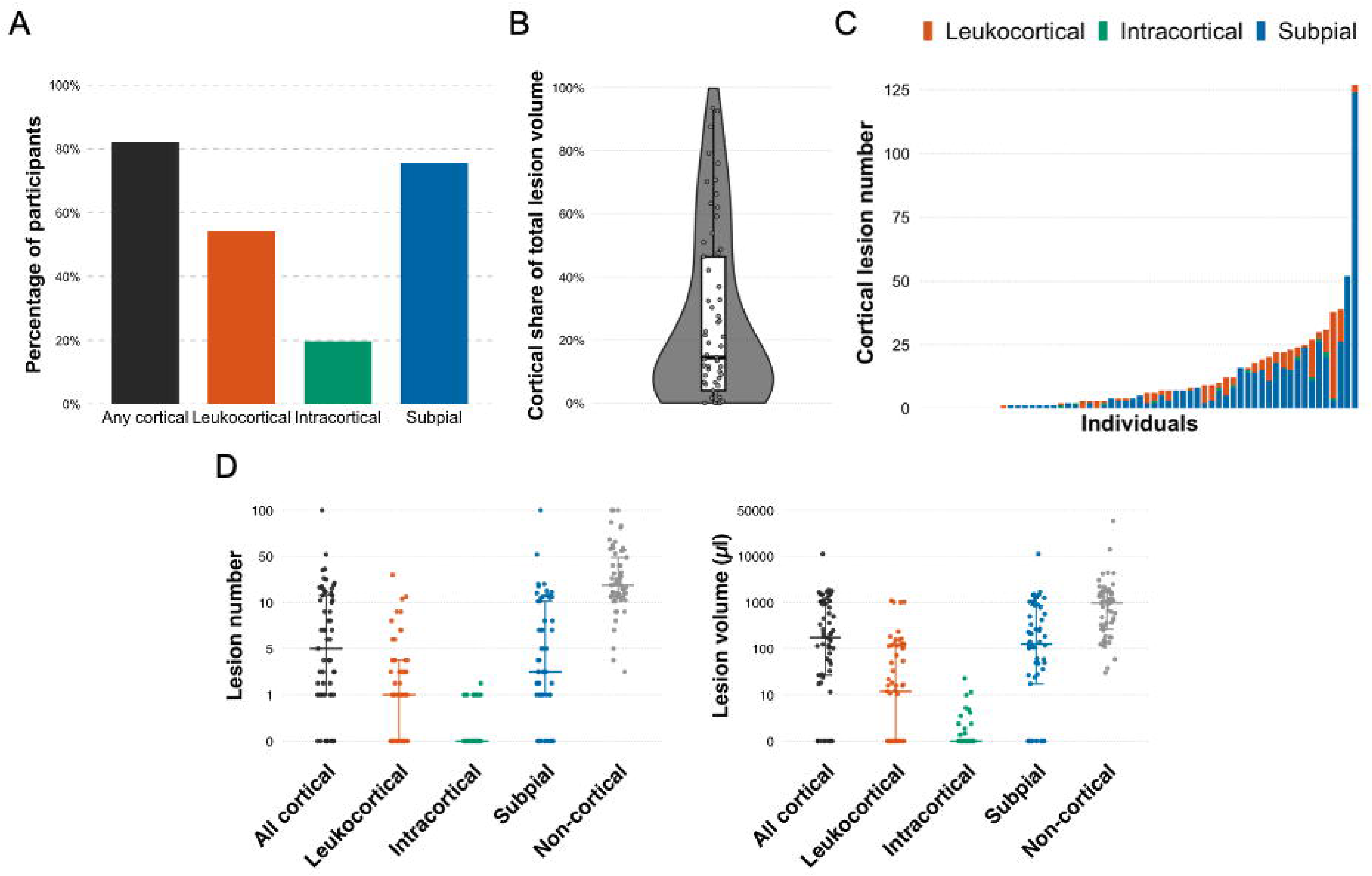
Cortical lesions are prevalent in newly diagnosed multiple sclerosis and, in many individuals, make up a substantial portion of brain lesion volume. **(A)** Percentage of participants in the newly diagnosed MS cohort with at least one lesion of the specified type. **(B)** Violin plot of the distribution of cortical share of total brain lesion volume. Hollow gray circles represent individual participants, bar represents median and interquartile range (Q1-3). **(C)** Per-participant cortical lesion counts, ordered by total count, with stacked colors indicating the number of each cortical lesion subtype. **(D)** Cortical and non-cortical lesion number and volume (µl). Each dot represents one participant. The thick horizontal line indicates the median, and the thinner horizontal lines and vertical bar span the interquartile range (Q1–Q3).

### The contribution of cortical lesions to total brain lesion burden is heterogeneous across individuals

The percentage of total brain lesion volume attributed to cortical lesions varied widely in the newly diagnosed MS cohort, with a median of 14% (IQR 43, range 0-94) (Figure 2B). In 13 participants (21%), cortical lesions constituted >50% of total lesion burden. Conversely, in 23 participants (38%), cortical fraction was less than 10%.

### Subpial lesions are the largest contributor to total cortical lesion volume in newly diagnosed multiple sclerosis

When categorizing cortical lesions by subtype, subpial lesions constituted the largest proportion of total cortical lesion volume (Figure 2C-D). The median subpial lesion volume was 192 µl (IQR 915, range 0-11970) compared to 17 µl (IQR 151, range 0-1366) for leukocortical lesions (p<0.001) and 0 µl (IQR 0, range 0-42) for intracortical lesions (p<0.001). The median percentage of cortical lesion volume classified as subpial was 87% (IQR 35, range 0-100), which was higher than the contribution of leukocortical (median 11%, IQR 40, range 0-100; p<0.001) or intracortical lesions (median 0%, IQR 0, range 0-24; p<0.001).

### Cortical and non-cortical lesion volumes are moderately correlated

Total cortical lesion burden was moderately correlated with non-cortical lesion burden (ρ=0.61, p<0.001 for number; ρ=0.39, p=0.002 for volume) (Figure 3). Non-cortical lesions were correlated with leukocortical lesions (ρ=0.62, p<0.001 for number; ρ=0.44, p<0.001 for volume) and subpial lesions (ρ=0.48, p<0.001 for number; ρ=0.34, p=0.006 for volume). In contrast, there was no association between intracortical lesions and non-cortical lesions (ρ=0.24, p=0.05 for number; ρ=0.14, p=0.25 for volume). Brain parenchymal volume was related to non-cortical lesion volume (ρ=-0.44, p<0.001; adjusted p=0.01), but not to cortical lesion volume (ρ=-0.16, p=0.24) (Figure 4).

**Figure 3.**
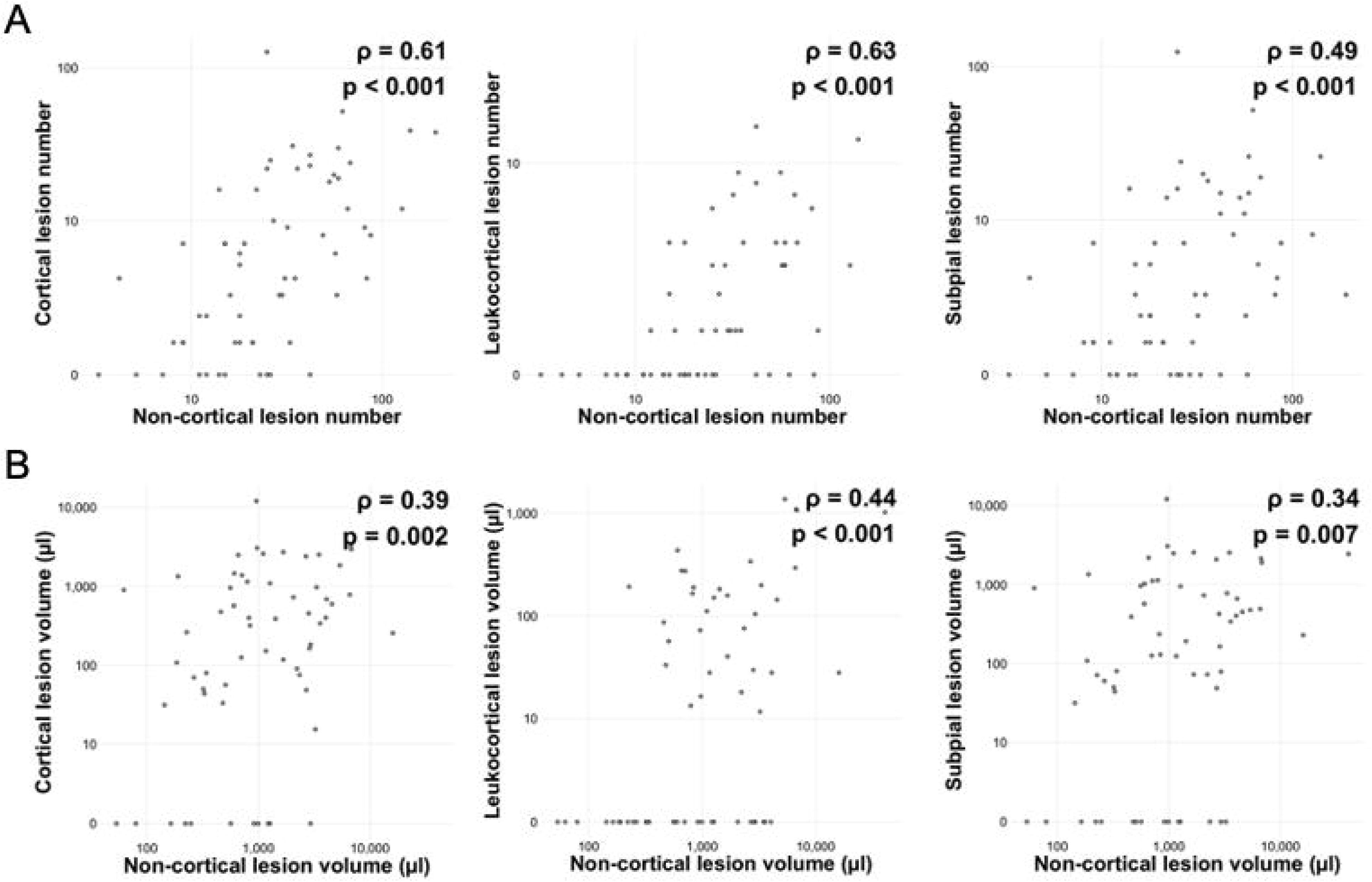
Cortical lesions moderately correlate with non-cortical lesions. **(A, B)** Scatter plots of non-cortical lesion number (A) and volume (B, µl) vs total cortical, leukocortical, and subpial lesion numbers and volumes (µl). Each dot represents one participant. Both axes are displayed on a pseudo-log_10_ scale. Spearman ρ and p-values are shown in each panel.

**Figure 4.**
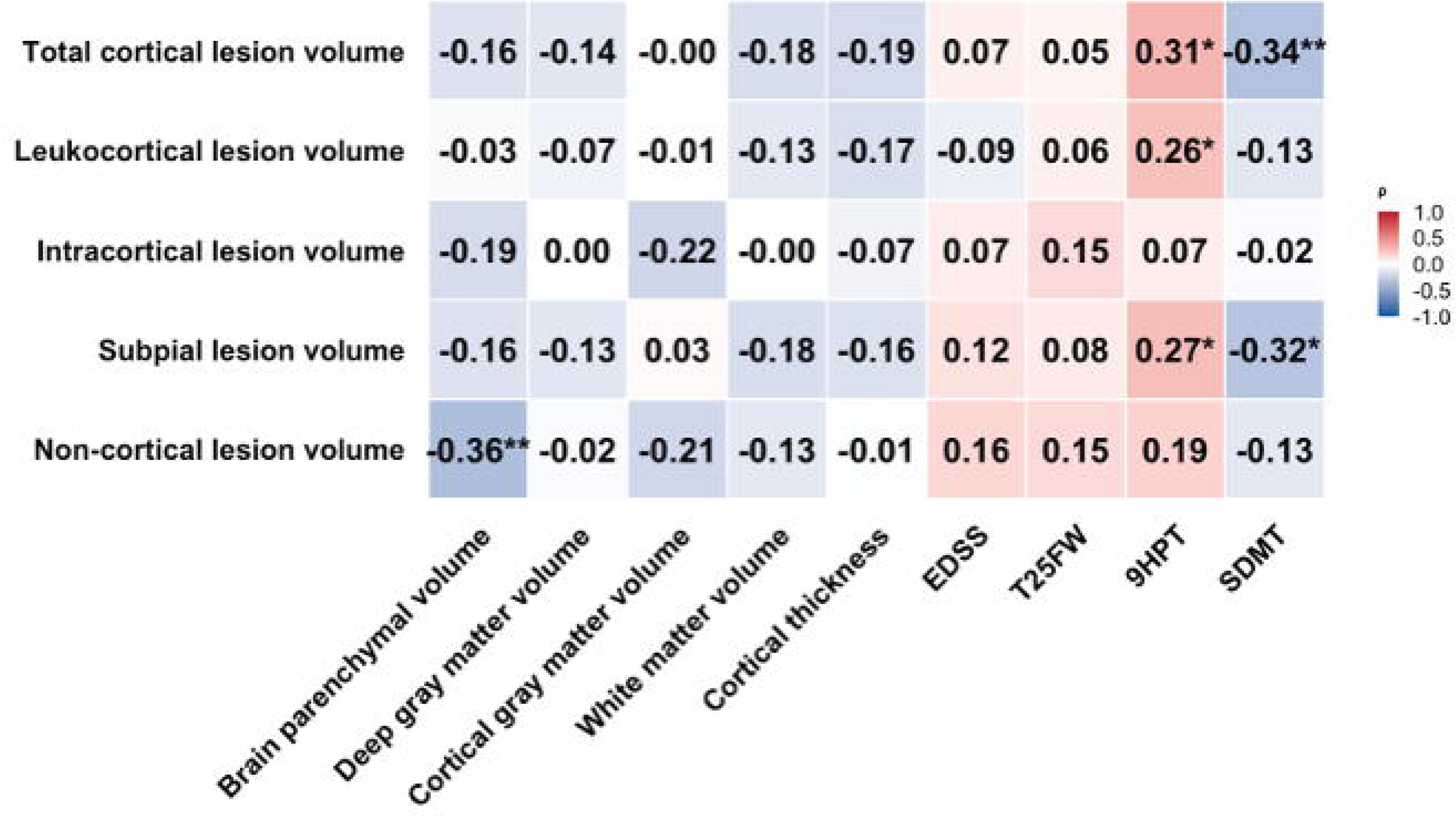
Cortical lesion volume is weakly associated with physical and cognitive disability in newly diagnosed multiple sclerosis. Heatmap displaying correlations between lesion volume and brain volumetric and clinical measures. Brain volumetric measures are normalized to the estimated total intracranial volume. Partial Spearman correlations were performed, adjusting for age. Color intensity reflects the magnitude of the correlation coefficient. Significance levels are indicated as follows: *: p<0.05, **: p<0.01, ***: p<0.001. After Benjamini-Hochberg false discovery rate correction, only the correlation between non-cortical lesion volume and brain parenchymal volume remained statistically significant (p=0.01). EDSS: Expanded Disability Status Scale. T25FW: Timed 25-foot walk. 9HPT: 9-Hole Peg Test (dominant hand). SDMT: Symbol Digit Modalities Test.

**Figure 5.**
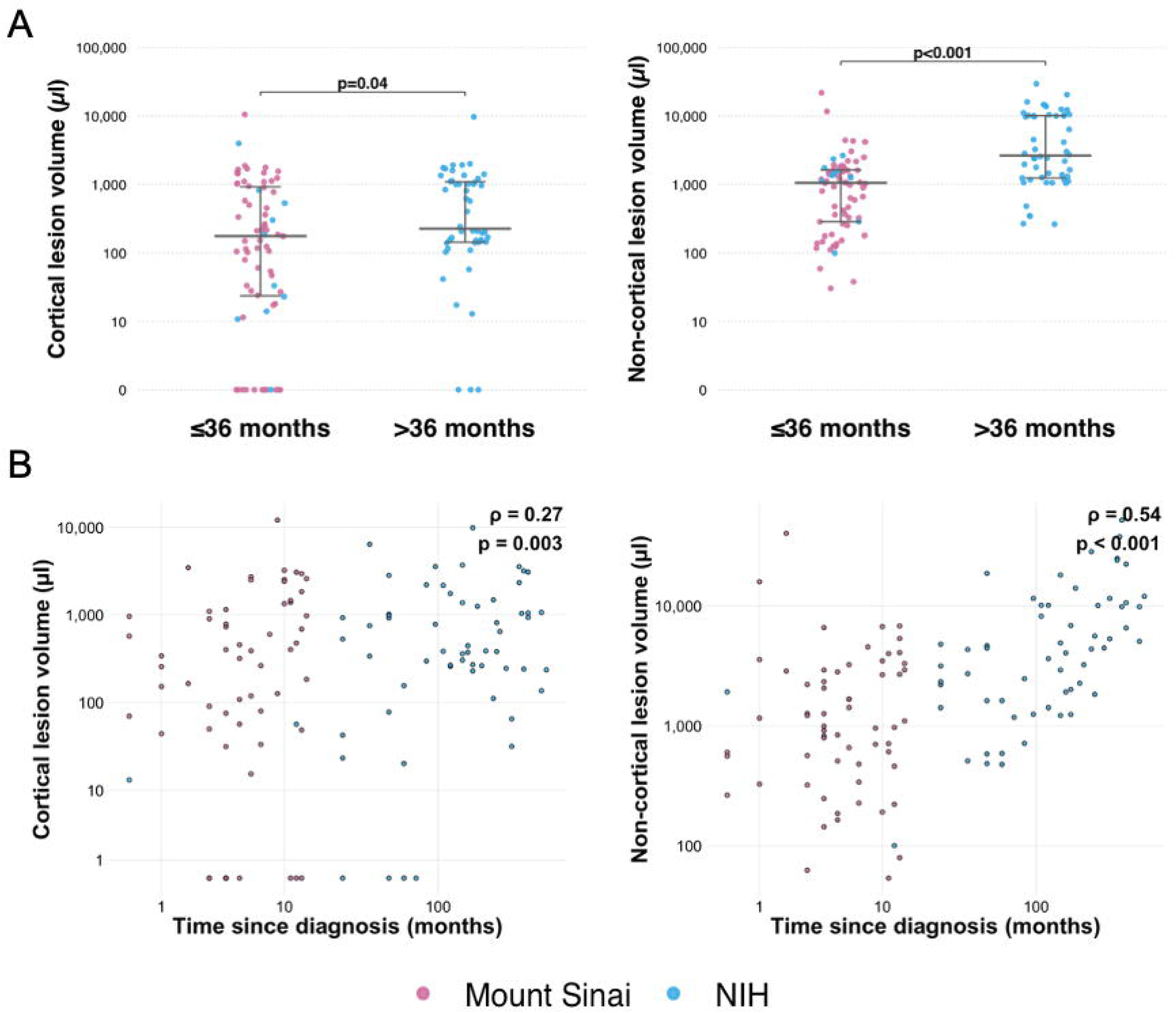
Cortical lesion volume is similar in early and established disease and is only weakly correlated with time since diagnosis. (A) Plots comparing cortical and non-cortical lesion volume in early (time since diagnosis ≤36 months) and established (>36 months) multiple sclerosis. The thick horizontal line indicates the median, and the thinner horizontal lines and vertical bar span the interquartile range (Q1–Q3). P-values from Mann-Whitney U tests are shown above each comparison. (B) Scatter plots showing Spearman correlations between time since diagnosis (months) and cortical and non-cortical lesion volumes. Cortical lesion volume was weakly associated with time since diagnosis, vs a moderate correlation between non-cortical lesion volume and time since diagnosis. Both the x- and y-axes are displayed on a pseudo-log_10_ scale. Correlation coefficients (ρ) and p-values are shown in each panel. Each dot represents one participant, with colors indicating the site of enrollment (Mount Sinai: purple; NIH: blue). NIH: National Institute of Health.

### Cortical lesion burden is weakly correlated with physical and cognitive disability in newly diagnosed multiple sclerosis

Cortical lesion number and volume were associated with worse performance on SDMT (ρ=-0.29, p=0.02 for number; ρ=-0.34, p=0.006 for volume) and dominant hand 9HPT (ρ=0.33, p=0.008 for number; ρ=0.28, p=0.02 for volume) (Figure 4, Supplementary Figure 1). These correlations were not significant after adjusting for multiple comparisons. There were no significant correlations between cortical lesions and EDSS or T25FW, and there were no correlations between non-cortical lesions and any of the disability measures.

### Cortical lesion volume is similar in early and established disease

To compare cortical lesion burden in early vs established disease, we pooled the Mount Sinai newly diagnosed MS cohort with a previously analyzed MS cohort from the NIH (n=60, median time since diagnosis 12 years, IQR 17, range 0-42). To determine consistency in lesion detection between sites, cortical lesions were identified by raters for the Mount Sinai cohort on a subset of 10 cases from the NIH cohort. Comparison of the consensus masks by both sets of raters showed excellent agreement in total cortical lesion counts, with an intraclass correlation coefficient of 0.974 (95% CI: 0.90-0.99), a Lin’s concordance correlation coefficient of 0.971 (95% CI: 0.90-0.99), and a Bland-Altman mean bias of -1.2 lesions (95% limits of agreement: -17.2 to 14.8). Comparison with the previous segmentations revealed no statistically significant difference in cortical lesion numbers (Mount Sinai cohort raters median 16, IQR 49, range 0-90 vs NIH raters with median 23, IQR 47, range 1-115; p=0.88).

Data from both cohorts were then pooled and divided into an early MS group (time since diagnosis ≤36 months, n=71, n=61 from the Mount Sinai cohort, n=10 from the NIH cohort) and an established MS group (time since diagnosis >36 months, n=50, all from the NIH cohort). The early MS group had a mean age of 36 ± 9 years and a median time since diagnosis of 0.5 years (IQR 0.7, range 0-3). In comparison, the established MS group was characterized by older age (mean 49 ± 11 years, p<0.001) and a greater median time since diagnosis (13.5 years, IQR 16.5, range 4-42; p<0.001) (Table 2).

**Table 2.**
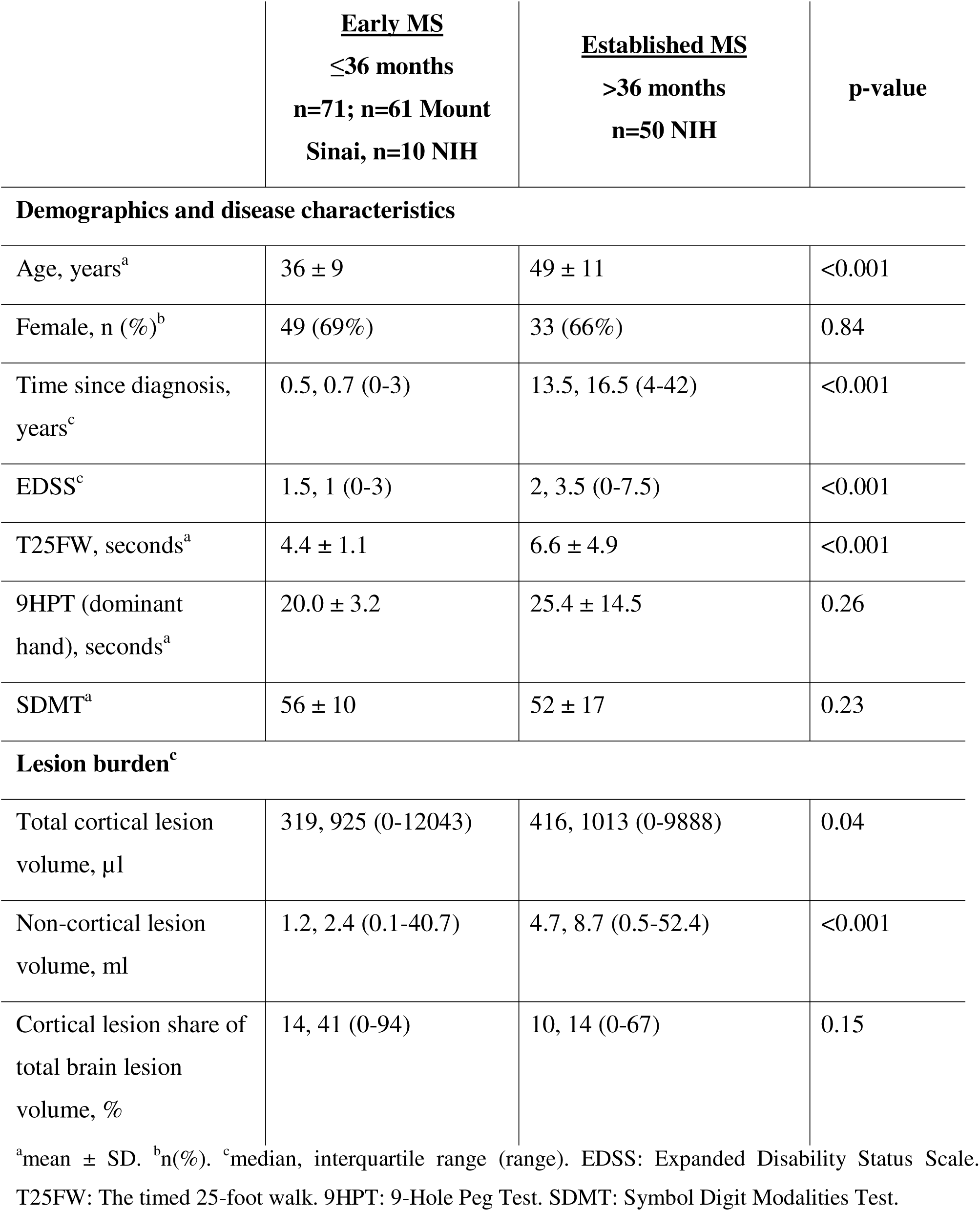
Demographics and disease characteristics of people with early (≤36 months since diagnosis) and established (>36 months since diagnosis) multiple sclerosis.

|  | **<u>Early MS</u>**<br>**$\leq 36$ months**<br>**n=71; n=61 Mount Sinai, n=10 NIH** | **<u>Established MS</u>**<br>**$> 36$ months**<br>**n=50 NIH** | **p-value** |
| --- | --- | --- | --- |
| **Demographics and disease characteristics** |  |  |  |
| Age, years<sup>a</sup> | 36 ± 9 | 49 ± 11 | <0.001 |
| Female, n (%)<sup>b</sup> | 49 (69%) | 33 (66%) | 0.84 |
| Time since diagnosis, years<sup>c</sup> | 0.5, 0.7 (0-3) | 13.5, 16.5 (4-42) | <0.001 |
| EDSS<sup>c</sup> | 1.5, 1 (0-3) | 2, 3.5 (0-7.5) | <0.001 |
| T25FW, seconds<sup>a</sup> | 4.4 ± 1.1 | 6.6 ± 4.9 | <0.001 |
| 9HPT (dominant hand), seconds<sup>a</sup> | 20.0 ± 3.2 | 25.4 ± 14.5 | 0.26 |
| SDMT<sup>a</sup> | 56 ± 10 | 52 ± 17 | 0.23 |
| **Lesion burden<sup>c</sup>** |  |  |  |
| Total cortical lesion volume, μl | 319, 925 (0-12043) | 416, 1013 (0-9888) | 0.04 |
| Non-cortical lesion volume, ml | 1.2, 2.4 (0.1-40.7) | 4.7, 8.7 (0.5-52.4) | <0.001 |
| Cortical lesion share of total brain lesion volume, % | 14, 41 (0-94) | 10, 14 (0-67) | 0.15 |

<sup>a</sup>mean ± SD. <sup>b</sup>n(%). <sup>c</sup>median, interquartile range (range). EDSS: Expanded Disability Status Scale. T25FW: The timed 25-foot walk. 9HPT: 9-Hole Peg Test. SDMT: Symbol Digit Modalities Test.

Non-cortical lesion volume was ∼3.5 times higher in the established MS group (median 4.7 ml, IQR 8.7, range 0.5-52.4) compared to the early MS group (median 1.2 ml, IQR 2.4, range 0.1-40.7; p<0.001). In comparison, total cortical lesion volume was only ∼1.3 times higher in the established group (median 416 µl, IQR 1013, range 0-9888) compared to the early group (median 319 µl, IQR 925, range 0-12043; p=0.04). Non-cortical lesion volume was moderately associated with time since diagnosis (ρ=0.54, p<0.001), as was cortical lesion number (ρ=0.50, p<0.001), whereas cortical lesion volume showed only a weak association (ρ=0.27, p<0.001), significantly weaker than that of non-cortical lesion volume (Steiger’s Z = 3.24, p=0.001).

## Discussion

Here, we characterize cortical lesions cross-sectionally in a newly diagnosed MS cohort using highly sensitive 7 T MRI. We find that most individuals with newly diagnosed MS have cortical lesions, with subpial lesions making up an average of almost 90% of cortical lesion volume. We also observe that in a substantial proportion of individuals with newly diagnosed MS (21% in our cohort), cortical lesions constitute over 50% of total brain lesion volume. Finally, we find that cortical lesion volume is ∼1.3 times greater in established vs early MS, despite ∼3.5 times greater non-cortical brain lesion volume in established MS. Taken together, these data suggest that cortical lesions predominantly form early in the MS disease course.

Current understanding of cortical pathology in MS derives largely from postmortem studies of individuals with longstanding, end-stage disease^2–4^, many of whom were likely never treated or treated with low-efficacy disease-modifying therapies for a relatively short period. The prevalence and extent of cortical lesions at the earliest stages of MS remain poorly characterized *in vivo*. The prevalence of cortical involvement we observed in early MS is consistent with a growing body of histopathological and imaging evidence suggesting that cortical demyelination begins at, near, or before symptom onset. Cortical demyelination was identified in biopsy samples from 53/138 people with MS (38%) undergoing diagnostic workup^1^. In addition, multiple studies using 7 T MRI have demonstrated the presence of cortical lesions in individuals within 3-5 years of diagnosis^24–26^. In a recent 7 T MRI study with a smaller cohort (n=20), cortical lesions were detected in 75% of individuals within one year of diagnosis^27^.

Our high-resolution *in vivo* data extend prior pathological and imaging observations by demonstrating that cortical lesion volume in early MS is comparable to that seen in established disease. Prior studies indicate that cortical lesion burden is higher, on average, in individuals with progressive vs relapsing disease^9,26^ and that individuals with higher baseline cortical lesion burden are more likely to experience worsening disability over time^9,13,28,29^. In combination with our data, this suggests that cortical lesion burden early in disease could be an important predictor of future disability worsening, which could be tested with long-term follow-up of our newly diagnosed cohort. The similarity in cortical lesion burden between early and longstanding disease is also consistent with a previous longitudinal study demonstrating a low rate of new cortical lesion formation^13^, although other longitudinal studies have reported higher rates, particularly in progressive MS^30,31^. These discrepancies likely reflect differences in cohort composition.

The effects of disease-modifying therapy on cortical lesion formation are unclear^32,33^, and it is possible that similar cortical lesion burden in early vs established MS demonstrated here reflects the effect of DMTs. MRIs for the NIH cohort were performed in 2017-2018, so similar lesion burden between early and established disease is unlikely to reflect the effects of high-efficacy disease-modifying therapy, but even lower efficacy DMTs could influence cortical lesion formation. However, the much greater increase in non-cortical vs cortical lesion volume between the early and established groups still suggests that cortical lesions form earlier than non-cortical lesions.

Among cortical lesion subtypes, subpial lesions were the dominant contributor to cortical lesion volume. This observation mirrors the subtype distribution reported in the earliest 7 T studies of MS cortical pathology as well as postmortem pathology data^3,8,34^. The proportion of subpial lesions in our cohort substantially exceeds that reported in prior 7 T studies using either T1w^35^ or T2*w^8^ images alone. Our combined use of high-resolution T1w and T2*w contrasts likely improves sensitivity to subpial pathology, further closing the gap between *in vivo* cortical lesion detection and neuropathological findings. The predominance of subpial lesions may be important due to their linkage in histopathological studies with meningeal inflammation and associated tissue injury, even in normal-appearing cortex, which could be a main driver of cortical pathology in progressive MS^4,36–38^. However, other studies have demonstrated little or no correspondence between meningeal inflammation and cortical demyelination^39^.

The capacity of high-resolution 7 T MRI to detect and classify cortical lesions is central to interpreting these findings. Even advanced 3T techniques, such as double inversion recovery^40^ and phase-sensitive inversion recovery^41^, detect a substantially smaller portion of the cortical lesions visible at ultra-high field strength and are particularly insensitive to subpial lesions^5^. We found that in approximately 20% of our newly diagnosed MS cohort, cortical lesions constituted over 50% of total brain lesion burden, which are largely undetected on standard clinical MRI. Thus, what has historically been classified as low or mild lesion burden in early MS in part reflects the insensitivity of conventional imaging rather than a truly indolent disease state, and individuals labeled as having “low lesion burden” may carry a less favorable long-term prognosis driven by largely invisible cortical pathology. These observations underscore the need for clinically feasible cortical lesion detection methods. Beyond diagnosis, the heterogeneity of cortical lesion burden across individuals in our cohort may also inform future trial design, with cortical lesion burden potentially serving as a stratifying biomarker in prospective studies.

Correlations between lesion burden and disability in our cohort were limited for both cortical and non-cortical lesions. This is not unexpected in an early MS cohort with low overall disability. Prior 7 T studies in patients with longer disease duration have consistently shown associations between cortical lesion burden and both cognitive and physical disability^26,35^. Longitudinal follow-up will be needed to determine whether early cortical lesion burden predicts subsequent clinical course.

Several limitations should be considered. Our comparison between early and established MS cohorts is cross-sectional rather than within-subject longitudinal, with early and established MS cohorts largely coming from two separate studies performed at different sites and with a five-year gap, which may confound comparisons. Although imaging methods for the two cohorts were very similar, MRIs were performed on different scanners with slightly different imaging protocols. In addition, our 0.5mm T1 and T2*w imaging does not fully cover the supratentorial brain, leading to potential underestimates of cortical lesion burden, and in a subset of cases from both cohorts, only 0.5 mm MP2RAGE images were available, leading to underestimation of total non-cortical lesion volume in these individuals.

In summary, this study provides evidence that cortical demyelination is already substantial and heterogeneous in early MS, and that subpial lesions are its primary component. The relatively modest increase in cortical lesion volume from early to established MS (∼1.3-fold), contrasted with the substantially greater accumulation of non-cortical lesion volume (∼3.5-fold), suggests that much of the cortical injury is established early, and this early window may determine long-term cognitive and functional trajectories. These findings argue for integrating MRI methods sensitive to cortical lesions into the evaluation of early MS, and they position cortical lesion burden as a candidate biomarker for risk stratification and clinical trials.

## Acknowledgements

We thank all participants for their time and contribution to this study. We thank the faculty and staff at the Corinne Goldsmith Dickinson MS Center at Mount Sinai and the Neuroimmunology Clinic at the NIH for their assistance with patient recruitment and study visits. We also thank the MRI technologists and imaging center staff for their technical support during data acquisition.

## Data availability

Deidentified data supporting the findings of this article will be made available upon publication.

