## Supplementary Figure 1 for "Cortical Lesions Form Predominantly in Early Multiple Sclerosis"

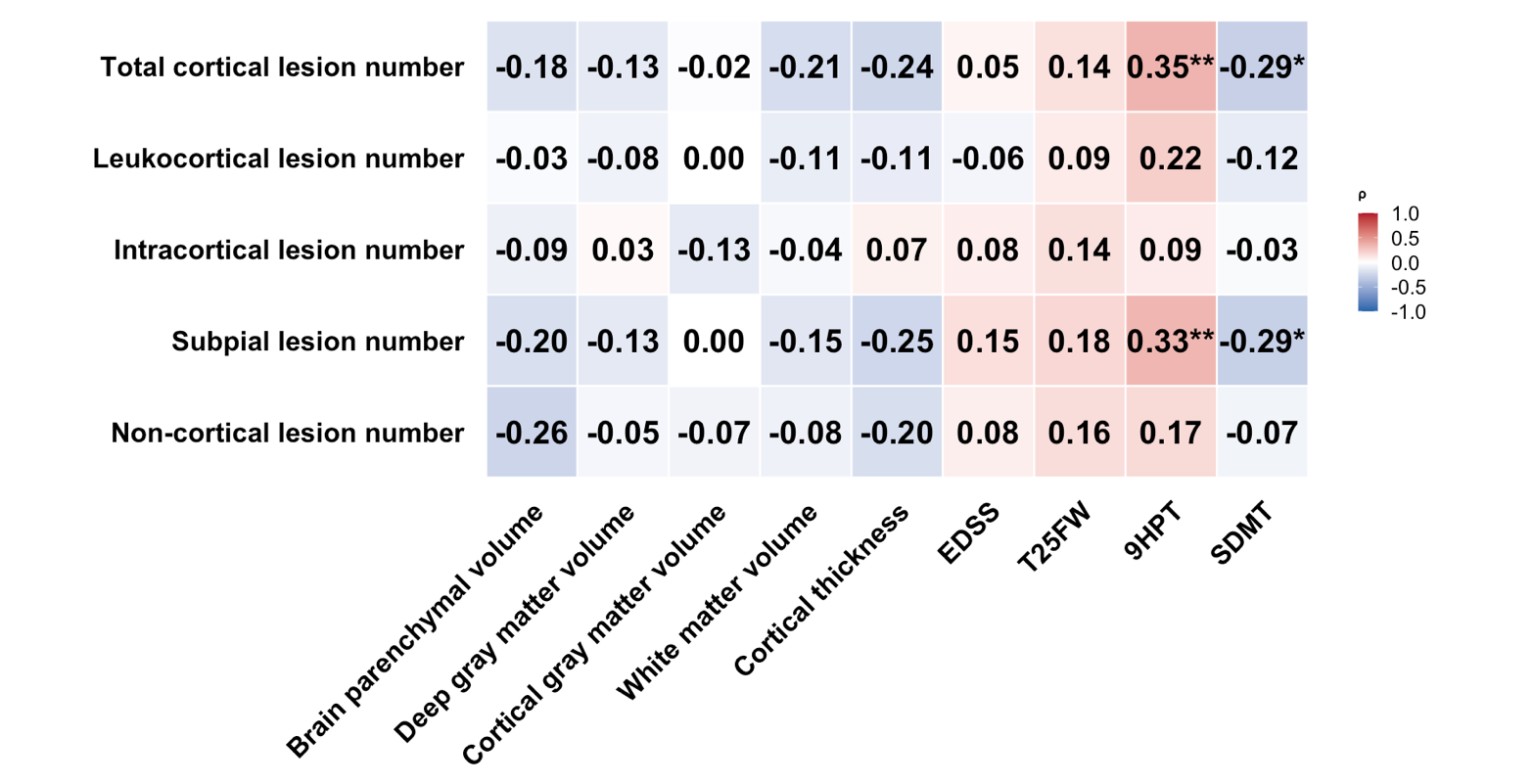

**Supplementary Figure 1. Cortical lesion volume is weakly associated with physical and cognitive disability in newly diagnosed multiple sclerosis.** Heatmap displaying correlations between lesion volumes and brain volumetric and clinical measures. Brain volumetric measures are normalized to the estimated total intracranial volume. Partial Spearman correlations were performed, adjusting for age. Color intensity reflects the magnitude of the correlation coefficient. Significance levels are indicated as follows: *: p<0.05, **: p<0.01. After Benjamini-Hochberg false discovery rate correction, any correlation remained significant (p>0.05). EDSS: Expanded Disability Status Scale. T25FW: Timed 25-foot walk. 9HPT: 9-Hole Peg Test. SDMT: Symbol Digit Modalities Test.
