## Supplementary Table 1 for "Cortical Lesions Form Predominantly in Early Multiple Sclerosis"

**Supplementary Table 1. MRI parameters**

| **7T Brain** | **Acquisition Plane** | **3D vs 2D** | **TR (ms)** | **TI (ms)** | **TE (ms)** | **FA (°)** | **AT (min:sec)** | **FOV (mm, AP x RL x SI)** | **In-plane resolution (mm)** | **Slice thickness (mm)** |
| --- | --- | --- | --- | --- | --- | --- | --- | --- | --- | --- |
| Mount Sinai | | | | | | | | | | |
| 0.5 mm MP2RAGE | Axial | 3D | 6000 | 1000, 2900 | 4.94 | 4, 5 | 10:41 | 168 x 224 x 112 | 0.5 x 0.5 | 0.5 |
| 0.7 mm MP2RAGE | Sagittal | 3D | 6000 | 800, 2700 | 3.09 | 4, 5 | 10:30 | 224 x 224 x 157 | 0.7 x 0.7 | 0.7 |
| Motion and B_0_ corrected T2*w GRE | Axial | 3D | 72.3 | NA | 12.26, 23.37, 34.48, 45.59 | 12 | 11:34 | 240x 180 x40 | 0.5 x 0.5 | 0.5 |
| NIH^1,2^ | | | | | | | | | | |
| 0.5mm MP2RAGE | Axial | 3D | 6000 | 1000, 2900 | 5.14 | 4, 5 | 10:32 | 224 x 168 x 112 | 0.5 x 0.5 | 0.5 |
| 0.7mm MP2RAGE | Sagittal | 3D | 6000 | 800, 2700 | 3.02 | 4, 5 | 10:08 | 224 x 157 x 224 | 0.7 x 0.7 | 0.7 |
| T_2_^*^w EPI | Sagittal | 3D | 52 | NA | 23 | 10 | 3:40 | 220 x 180 x 88 | 0.5 x 0.5 | 0.5 |
| T_2_^*^w GRE | Axial | 2D | 4095 | NA | 11.4, 22.5, 33.6, 44.7, 55.8 | 90 | 11:26 | 240 x 168 x 30 | 0.5 x 0.5 | 0.5 |
| Motion and B_0_ corrected T_2_^*^w GRE | Axial | 3D | 74 | NA | 18, 29.5, 41.0, 52.4 | 10 | 11:50 | 240x180x32 | 0.5 x 0.5 | 0.5 |

7T whole-body research system (Siemens, Erlangen, Germany) with a single-channel transmit, 32-channel phased array receive head coil. Inversion pulse type was recorded for each session and accounted for in subsequent analyses.

TR: repetition time. TI: inversion time. TE: echo time. FA: flip angle. AT: acquisition time. FOV: field of view. AP: anterior-posterior. RL: right-left. SI: superior-inferior. MP2RAGE: magnetization prepared 2 rapid gradient echoes. T2*w GRE: T2* weighted gradient recalled echo. T2*w EPI: T2* weighted echo-planar imaging.
